# Spatiotemporal droplet dispersion measurements demonstrate face masks reduce risks from singing: results from the COvid aNd FacEmaSkS Study (CONFESS)

**DOI:** 10.1101/2021.07.09.21260247

**Authors:** Kai Man Alexander Ho, Hywel Davies, Ruth Epstein, Paul Bassett, Áine Hogan, Yusuf Kabir, John Rubin, Gee Yen Shin, Jonathan P. Reid, Ryo Torii, Manish K. Tiwari, Ramanarayanan Balachandran, Laurence B. Lovat

**Affiliations:** WEISS Centre, Division of Surgery & Interventional Science, University College London, Charles Bell House, 43-45 Foley Street, London, W1W 7TY; Department of Mechanical Engineering, University College London, Malet Place, London, WC1E 6BT; Department of Otolaryngology, Royal National Ear Nose and Throat & Eastman Dental Hospital, University College London Hospitals NHS Foundation Trust, 47-49 Huntley Street, London, WC1E 6DG; Statsconsultancy Ltd, 40 Longwood Lane, Amersham, Bucks, HP7 9EN; Department of Virology, University College London Hospitals NHS Foundation Trust, 250 Euston Road, London, NW1 2PG; School of Chemistry, University of Bristol, Cantock’s Close, Bristol, BS8 1TS

## Abstract

**Background:** COVID-19 has restricted singing in communal worship. We sought to understand variations in droplet transmission and the impact of wearing face masks.

**Methods:** Using rapid laser planar imaging, we measured droplets while participants exhaled, said ‘hello’ or ‘snake’, sang a note or ‘Happy Birthday’, with and without surgical face masks. We measured mean velocity magnitude (MVM), time averaged droplet number (TADN) and maximum droplet number (MDN). Multilevel regression models were used.

**Results:** In 20 participants, sound intensity was 71 Decibels (dB) for speaking and 85 dB for singing (p<0.001). MVM was similar for all tasks with no clear hierarchy between vocal tasks or people and >85% reduction wearing face masks. Droplet transmission varied widely, particularly for singing. Masks decreased TADN by 99% (p<0.001) and MDN by 98% (p<0.001) for singing and 86-97% for other tasks. Masks reduced variance by up to 48%. When wearing a mask, neither singing task transmitted more droplets than exhaling.

**Conclusions:** Wide variation exists for droplet production. This significantly reduced when wearing face masks. Singing during religious worship wearing a face mask appears as safe as exhaling or talking. This has implications for UK public health guidance during the COVID-19 pandemic.

## Introduction

Airborne transmission of SARS CoV-2 is now known to be the dominant pathway for disease spread in the COVID-19 pandemic.^1^ Both Public Health England (PHE) and Centers for Disease Control and Prevention (CDC) in the United States have recommended social distancing, wearing face coverings and avoiding large gatherings to reduce viral transmission risk.^2,3^ The risks of large gatherings were highlighted early on by well publicised examples such as the Shincheonji Church of Jesus in South Korea and the Sri Petaling mass gathering in Malaysia which accounted for over 60% and 35% of cases in their respective countries at their peaks.^4,5^ Group singing, which is often an integral part of religious worship, has also been linked to clusters of infection, most notably the Skagit County choir cluster in which up to 53 of 61 participants were infected from a single index case.^6^

Restrictions have been placed on indoor singing: guidance issued by PHE on the principles of safe singing state that singing should only take place in larger well-ventilated spaces or outdoors with social distancing applied.^7^ In practice, United Kingdom guidance has prohibited but not outlawed indoor singing since July 2020, with outdoor singing returning only in April 2021.^8,9^ This has had a major impact on communal religious worship. Participants obtain great spiritual elevation from song but they also follow guidelines closely and have suffered greatly as a result.^10^

Expiratory events such as talking, coughing and sneezing generate a plume of respiratory particulates which vary in size.^11,12^ These range from muco-salivary droplets originating from the oral cavity and pharynx to microscopic aerosols from the small airways of the lungs.^11–13^ SARS-CoV-2 can be transmitted both via respirable aerosols, normally defined as droplets with an initial size <5-10 µm in diameter that rapidly dehydrate and can remain suspended for hours (often designated as airborne transmission), and larger droplets.^14^ The range of droplet transport is dependent on their size: droplets with initial diameter >100 µm settle onto nearby surfaces, while ‘coarse aerosols’ comprising droplets that are <100 μ m when produced, dehydrate, remain suspended for prolonged periods and travel long distances in the air.^15^ They are therefore aerodynamically similar to respirable aerosols, particularly once their reduction in size due to evaporation of water on exhalation from the humid respiratory tract is accounted for.^14–16^ In terms of viral transmission risk, most expiratory droplets are less than 1 μ m in diameter, but speech and singing both produce additional particles which peak in size at 3.5-5 μ m.^12,13,17^ Droplets with initial diameters of less than 1-3 μ m are unlikely to contain significant viral load when compared to the coarser sized particles produced during vocalisation.^18,19^ In contrast, surgical face masks can successfully block shedding of coronavirus and other seasonal viruses where the droplet particles are more than 5 μ m in diameter.^20^ Thus, quantifying droplet generation and reducing transmission in the 1-5 μ m range is likely to be of high importance. In this paper, the term droplets will be used generically and refer to both respirable and coarse liquid aerosol particles. Droplets may dehydrate to create solid particles.

Several factors make singing higher risk for transmission of SARS-CoV-2 and other airborne viruses compared to normal speech. These include higher frequencies, continuous voicing and more articulated consonants.^21^ Mitigation factors have been studied to make singing safer. Echternach et al. investigated dispersion dynamics of aerosols in 10 professional singers and recommended up to 2.5 m social distance to persons in front and 1.5 m to the side to reduce aerosol droplet spread.^22^ Loud singing with a face mask reduced the number of aerosol droplets to a level similar to normal talking, although this was not statistically significant.^21^ Interestingly, PHE guidelines suggest that the evidence for face coverings in singing is uncertain, but do state ‘their use might be considered as additional precautionary mitigation, where this is practicable.’^7^ Single use surgical face masks can capture coarse and fine respiratory aerosol droplets of sizes as small as 1-5 μ m^20^ and it is possible to make reusable face masks with similar efficacy.^18^

Planar laser imaging (PLI) can capture images of droplet transmission at speeds of up to 3000 frames per second (fps). Previous studies of respiratory droplet transmission have either examined coughing,^23^ have been undertaken at low time resolution (such as 125 fps^21^) or have not demonstrated statistically significant differences between people in the spatio-temporal evolution of droplets when a verbal task is performed.^21^ This is obviously important for understanding disease transmission risks. Furthermore, such high-speed imaging techniques have not been exploited to thoroughly investigate the effect of wearing a face mask during singing to the best of our knowledge. We sought to explore all these issues in our study, using PLI with high-speed image capture in a large cohort of volunteers while speaking or singing with and without face masks.^24,25^

## Aims

The aims were to investigate the differences in droplet transmission between a variety of vocal tasks including singing, and to offer insights on the effect of wearing a face mask on droplet transmission.

There were four specific objectives:

1. To understand the differences in droplet transmission between different vocal tasks.
2. To examine inter-participant variability, with particular interest in singing.
3. To explore the relative difference in the number of droplets for singing compared to speaking and exhaling when wearing a face mask.
4. Finally, to explore the concept of ‘Super Emitters’ by analysing whether any participants transmitted many more droplets than others for individual tasks.

## Methods

### Study Population

The CONFESS study (COvid aNd FacE maSkS) was designed as a cross-sectional study to assess the safety of singing in religious worship during the COVID-19 pandemic. Participation was voluntary. Participants were recruited via social media, traditional media outlets including BBC News and targeting of religious groups.^26^ Participants consented and enrolled in the study on the dedicated website (https://www.confess-study.co.uk/) and completed a study questionnaire between September and October 2020. A subgroup of 20 people were invited to participate in experiments at the Mechanical Engineering Department in University College London, London, UK. This group was chosen to represent a wide range of demographics including age, sex, racial background and body habitus. Experiments took place in October 2020 but were curtailed when the second UK-wide lockdown was announced at three-days’ notice.^27^

### Experimental Design

Participants completed vocal tasks in a specifically designed apparatus (Figure 1 and Supplementary Figure 1). Institutional and nationally mandated COVID-19 and laser safety precautions were taken. Vocal tasks were completed without a face mask and subsequently while wearing a type IIR surgical face mask (OPROtec, Hemel Hempstead, UK). In all cases only a single performance of each task was recorded per participant (e.g., a single utterance of ‘hello’), due to time constraints. These tasks were:

i. Exhaling normally
ii. Saying ‘hello’
iii. Saying ‘snake’
iv. Singing the note ‘la’
v. Singing the first two lines of the song ‘Happy Birthday’

**Figure 1:**
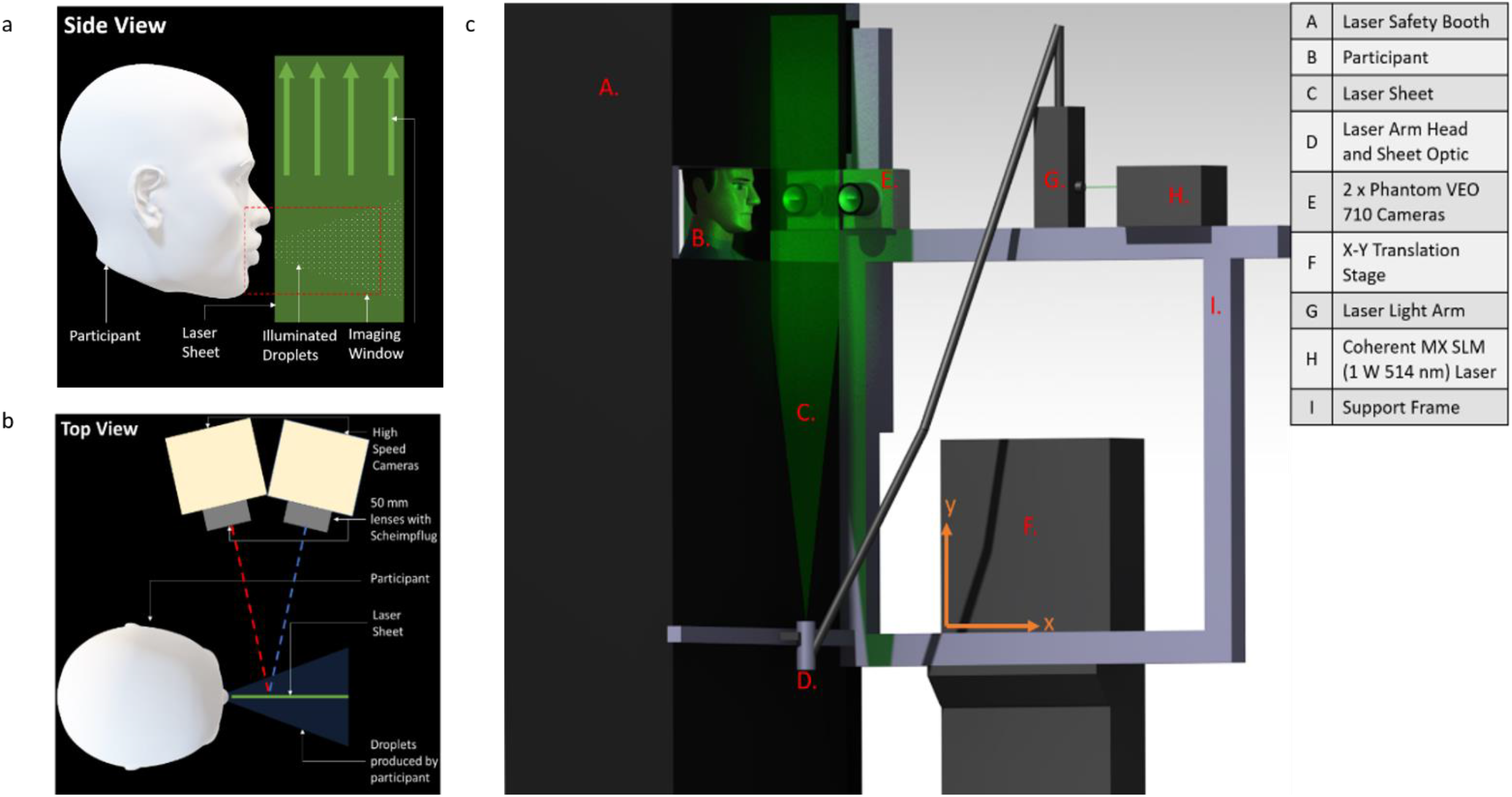
Diagram demonstrating a) side view and b) top view of laser sheet and camera alignment. Panel c) demonstrates the custom rig built for the experiment, including a laser safety booth for the participant.

These words were chosen as they included a combination of voiceless fricatives such as /h/ and /s/, which are consonants produced by forcing air through a channel created by certain movements of the articulators such as lips, tongue, teeth or palate as well as the voiced consonants /p/ and /b/ which are produced with vibration in the vocal folds.^23^ To simulate real-life, participants were instructed to say the words as if speaking to somebody in a real conversation and to sing in their typical voice. They were specifically told ‘to sing in a key and volume you find pleasurable and comfortable’. A sound level meter (Sauter SU130, Balingen, Germany) was placed 300 mm forward of the participant’s mouth to capture the far field sound intensity.

### Droplet Detection and Validation

Droplets were detected using a laser planar illuminated imaging technique, PLI (Figure 1). In brief, participants stood in a laser safe booth with a head brace to minimise movement (Supplementary Figure 1). A 0.5 mm thick laser sheet was produced in front of the participant transverse to the exhaled air flow direction using Coherent MX SLM (1 W 514 nm) continuous laser with LASERPULSE^™^ light arm and sheet optics (TSI, Shoreview, Minnesota, USA). Two Phantom VEO 710 cameras (1280 x 800 pixels) (Vision Research, Wayne, New Jersey, USA) fitted with NIKKOR 50 mm lenses (Nikon, Tokyo, Japan) were used to capture high-speed photographs of droplets as participants completed the vocal tasks. To avoid the laser shining directly onto the participants’ faces, a 25-30 mm gap in front of the mouth was not illuminated (Figure 1a). Images were captured for 2 seconds at 3000 frames per second (fps) for all tasks except singing ‘Happy Birthday’ where the frame rate was 1000 fps as the expected increased task duration would require greater data storage capacity which was constrained. Exposure time was kept constant for all tasks at 0.3 milliseconds.

We performed a validation to ensure that the laser could illuminate the droplets generated by a nebuliser, which are typically 0.3-4 µm, as measured by an aerodynamic particle sizer.^11^ We used an Omron NE-C28P medical compressor nebuliser (Omron, Kyoto, Japan), which produces droplets with a mass median aerodynamic diameter (MMAD) of 3 µm. Our imaging system readily detected the emitted droplets from the nebuliser. Therefore, we are confident we can detect particles of *at least* 3 µm, i.e. within the respiratory aerosol mode, although a precise lower limit of detection is hard to define.^12,21^

### Image Analyses

Once images were captured, they were entered into an in-house detection script written in MATLAB software (MathWorks, Natick MA, USA); this provided positional and spatial concentrations of illuminated droplets. Droplets were imaged in an image window of 170 mm x 110 mm x 0.5 mm, which was fixed for all participants and tasks. In all cases measurements were taken from an analysis window which was a 25 x 25 mm area located 35-60 mm from the mouth. We employed Particle Tracking Velocimetry (PTV) to track individual droplets using the open-source package TracTrac for MATLAB. With the described optical setup, the image pixel resolution was 135 µm.

We also used INSIGHT 4G Particle Imaging Velocimetry (PIV) software (TSI, Shoreview, Minnesota, USA). This measured the distance travelled by an individual droplet across two serial images. PTV accuracy is dependent on a higher number (100s) of images containing significant number of particles. For PIV the concentration of droplets must be higher for valid measurements to be attained.

We used the following measures to quantify both the numbers and velocity of droplets detected when participants were performing vocal tasks:

- Mean Velocity Magnitude (MVM): This provides a measure of the speed of droplets at a fixed point of 30 mm from the mouth, with droplets having transited through a mask if worn, using both PTV and PIV methods during the most significant vocal event, which was defined as longest sequence of images with the highest total number of droplets. Images were only analysed when droplets were detected for at least 200 continuous frames (approximately 0.067 seconds) to ensure the PTV tracking software did not generate spurious results.
- Time averaged droplet number (TADN): the sum of droplets over the duration of the task divided by the time taken. Significant portions of each recording showed no droplet transmission; these time portions were removed to improve statistical accuracy. Where minimal droplets were exhaled (e.g. participants 2 and 5 for ‘snake’ in Figure 2B), an arbitrary minimum of 500 milliseconds was chosen to ensure the entire task was assessed, and where a face mask was worn with no transmission of droplets, the identical time segment was chosen to compare droplet transmission between wearing and not wearing the mask.
- Maximum Droplet Number (MDN): This gives an indication of peak concentration of droplets by measuring the number of droplets per frame in the 30 consecutive frames with the highest droplet numbers.

**Figure 2:**
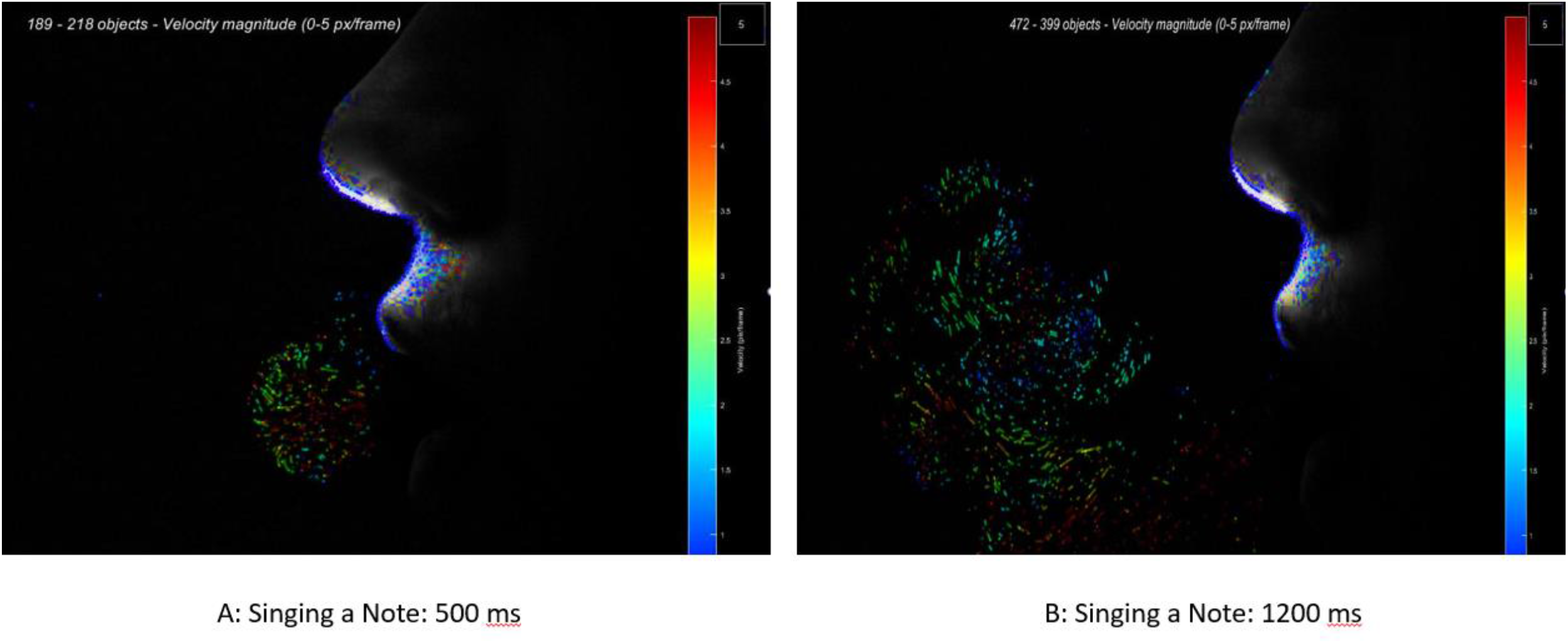
Panel A: Representative images showing time vector plots of droplets being produced. Colours show velocity and arrows show direction of movement of each individual droplet. These images are taken from 500 miliseconds and 1200 miliseconds after one of the participants sang the note ‘la’. These data were analysed to create the graphs in panel B. A full video sequence is available in supplementary materials.

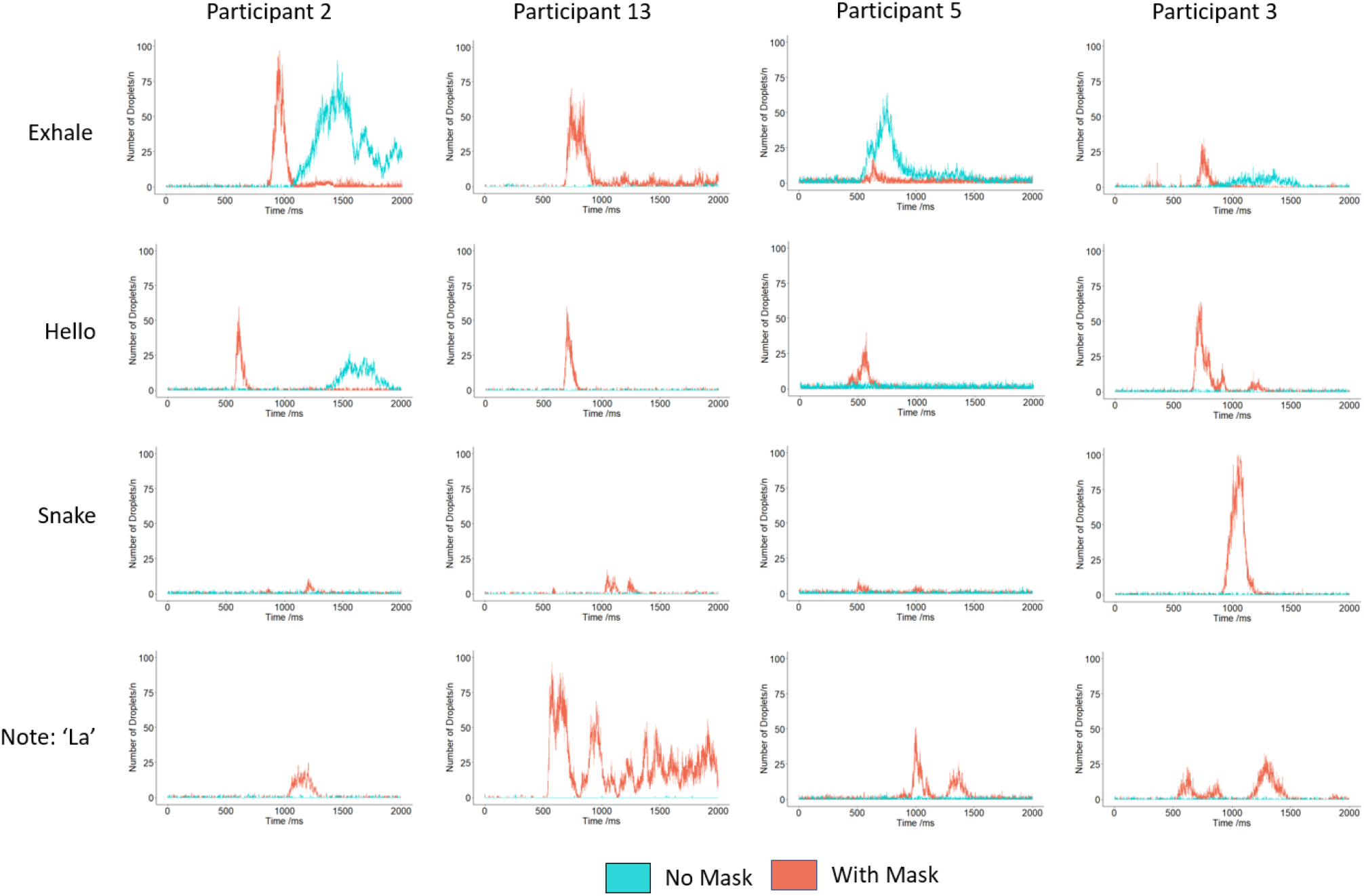
B: Representative time resolved transmission of droplets. There was no consistent pattern for numbers of droplets produced across the various tasks. Leakage through the mask occurred almost exclusively during exhaling. Otherwise, virtually no droplets were transmitted for any task when wearing masks.

### Data Analyses

Results and statistical tests were performed using R software 4.0.4 (R Foundation for Statistical Computing, Vienna, Austria). We used descriptive statistics for demographic data. Normally distributed data, such as sound intensity was evaluated using Student’s t-tests.

Data was collected serially over time for each participant both with and without a mask, giving multiple droplet measurements for each subject. As no droplets were observed in a large proportion of individual measurements, the data was heavily skewed and could not be transformed to a normally distributed scale. As a result, the analysis approach used for TADN was to consider the number of droplets as a count variable. The data was assumed to follow distributions commonly used for count data, specifically the negative binomial distribution. This was preferred to the Poisson distribution, also used for this type of data, due to the large variation in counts.

To allow for the repeat measurements from the same subjects, both over time, and with/without a mask, all analysis was performed using multilevel (mixed) regression models. Two level models were used with individual droplet measurements nested within participants. Specifically, multilevel negative binomial regression was used for the analyses.

For TADN, median and inter-quartile range (IQR) were analysed using the Wilcoxon signed-rank test.

### Ethical Approval

Ethical approval for the study was obtained from University College London Ethics Committee (Approval: 14223/002).

## Results

### Demographics

Twenty participants were included in the final analysis; 13 performed a sequence of 4 tasks: exhaling normally, saying ‘hello’ and saying ‘snake’ and singing a note. The remaining 7 participants sang the first two lines of the song ‘Happy Birthday’. There was inadequate time to perform the other tasks due to the prolonged data storage process after each task and the impending imposition of the second UK lockdown. This necessitated faster turnaround between participants than was ideal. Two participants accidentally knocked the laser apparatus during a single task. We have excluded results from these tasks in our analyses. Participants had a median age of 42.0 (interquartile range (IQR) = 27.0). 14/20 (70%) were female (Supplementary Table 1).

### Vocal tasks

Figure 2B illustrates the wide variation in droplet-time profiles of different vocal tasks. Data from all participants are shown in Supplementary Figure 2. Mean (+/- Standard Deviation (SD)) sound intensity for spoken tasks was 71 (+/-5.3) dB and 85 (+/-7.4) dB for singing (p<0.001). Mask wearing did not affect sound intensity (p=N.S.). Typically, ‘hello’ yielded a sharp peak associated with ‘h’, while ‘snake’ had 2 smaller peaks associated with ‘s’ and ‘k’. Singing a note led to droplet transmission over a longer period compared to speaking either ‘hello’ or ‘snake’ and participants transmitted more droplets. The variation between the participants is seen clearly as is the egress of air in a small number of participants when exhaling or saying ‘hello’ whilst wearing a mask.

### Mean Velocity Magnitude (MVM)

MVM was measured for each participant for each task. Without face masks, median MVMs were similar across all vocal tasks, ranging from 0.47 m/s for singing a note to 0.78 m/s for exhaling (Figure 3). When wearing masks, for most participants, there was no leakage at all. There was leakage for 2 participants when exhaling and 1 saying ‘hello’. In these cases, the MVM was reduced by 85% to 0.19 m/s from 1.28m/s for exhaling with a mask and to 0.14 m/s from 0.91 m/s when saying ‘hello’.

**Figure 3:**
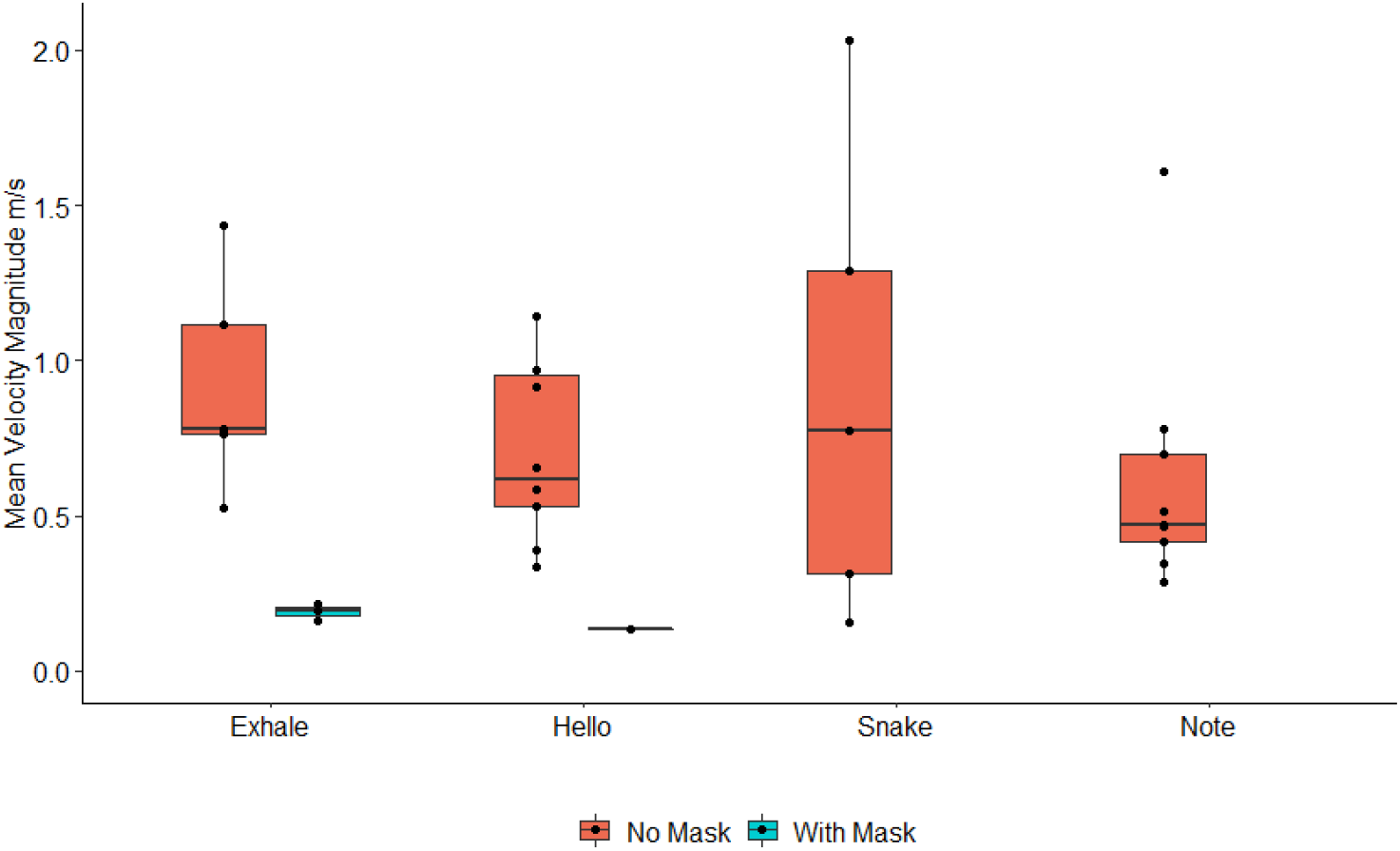
Boxplot with individual data points for mean velocity magnitude (MVM) across different vocal tasks. PTV accuracy is dependent on a higher number (100s) of images containing significant number of particles. For PIV the concentration of droplets must be higher for valid measurements to be attained. Several participants, for some tasks did not produce droplets to a level that satisfies the above conditions, as such no meaningful measurements could be taken. Data are not available for saying ‘snake’ with a mask, singing a note with a mask and singing ‘Happy Birthday’.

### Time Average and Maximum Droplet Number (TADN and MDN)

Table 1 shows the TADN emitted for each task. Although not normally distributed, the mean was the preferred summary measure as it better represents a measure of the total droplets compared to the median value. The TADN was lowest for saying ‘snake’, then exhaling, then saying ‘hello’ and then singing a note. Of interest, singing ‘Happy Birthday’ generated a lower TADN than saying ‘hello’ or singing a note. Figure 4 shows MDNs which demonstrated the same pattern. There was a statistically significant reduction in TADN of 86-99% for all tasks except exhaling where there were several cases of droplet transmission through the masks where TADN fell by 53%. The reductions were statistically significant for all tasks (Table 1). As droplets were travelling more slowly and were therefore visible in more frames, the actual number of droplets was probably even lower. Similarly, the MDN was significantly reduced by 86% for saying ‘snake’, 96% for saying ‘hello’ and 98% for singing a note. There was a trend to reduction for exhaling (86%) and singing ‘Happy Birthday’ (94%) which did not reach statistical significance (Figure 4).

**Table 1:** Comparison of time averaged droplet number with and without a face mask

| Task | No Mask<br>Mean ± SD | Mask<br>Mean ± SD | Ratio (*)<br>(95% CI) | Fold-Reduction<br>(95% CI) | P-value |
| --- | --- | --- | --- | --- | --- |
| Exhale (‡) | 3.80 ± 10.07 | 5.07 ± 11.84 | 0.461 (0.447, 0.475) | 2.17 (2.11, 2.24) | <0.001 |
| Hello | 8.38 ± 17.36 | 1.49 ± 3.42 | 0.109 (0.105, 0.114) | 9.2 (8.8, 9.5) | <0.001 |
| Snake | 3.56 ± 11.90 | 0.22 ± 0.49 | 0.113 (0.108, 0.118) | 8.8 (8.5, 9.3) | <0.001 |
| Note | 13.56 ±<br>17.82 | 0.12 ± 0.38 | 0.011 (0.010, 0.011) | 92.7 (89.2, 96.4) | <0.001 |
| Happy<br>Birthday | 6.58 ± 17.68 | 0.63 ± 1.39 | 0.142 (0.130, 0.155) | 7.0 (6.5, 7.7) | <0.001 |

(\*) Odds ratio expressed as number of droplets using mask relative to no mask
(‡) In a small number of participants, significant air egress through the mask led to a higher mean time averaged droplet number with a mask than without. Overall, TADN fell by more than 50% when wearing a mask

**Figure 4:**
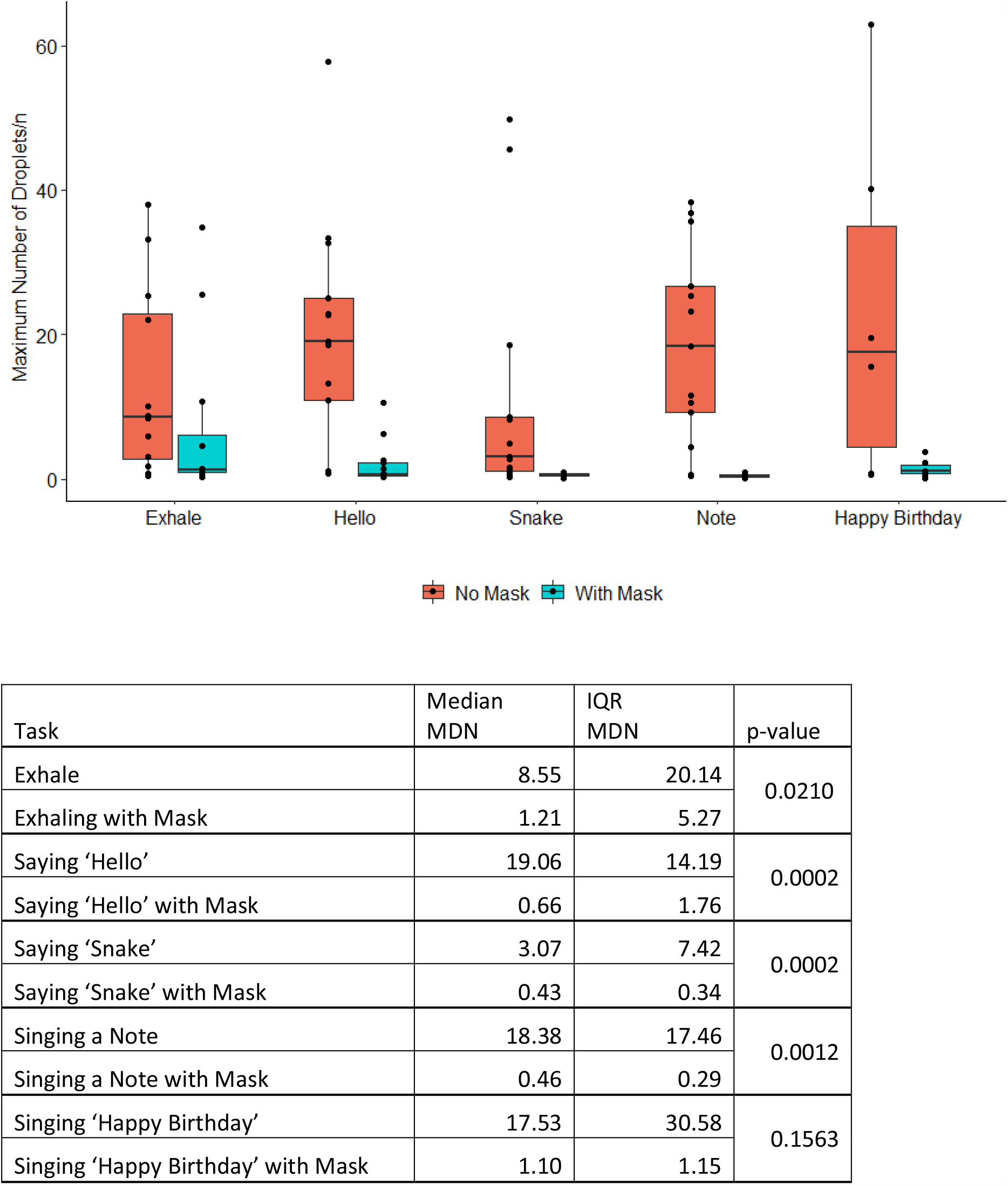
Boxplot demonstrating maximum droplet number (MDN) across a number of vocal tasks. Sub-table presents median and interquartile range (IQR) of each task, with Wilcoxon signed-rank tests to compare each vocal task with and without a face mask.

### Inter Participant Variability

We examined the inter-participant variability, with particular interest in singing a note as this generated the largest number of droplets. Multilevel regression analyses were used to extract the between-subject variance when a face covering was and was not used. A summary of these components of variance is presented in Table 2. The raw values are not particularly interpretable, with the focus on the relative size of the variation in one situation compared to the other. Inter-subject variance was considerably lower when a mask was worn compared to when a mask was not worn. Face masks effectively abolished the difference in transmission of droplets between high and low emitters.

**Table 2:** Variance in droplet production between participants when singing a note and ‘Happy Birthday’

| Task | Condition | Between-participant variance (*)<br>($\times 10^{-3}$ scale)<br>Estimate (95% CI) |
| --- | --- | --- |
| Happy Birthday | No mask | 2.85 (1.31, 6.14) |
|  | Mask | 1.47 (0.68, 3.19) |
| Note | No mask | 2.58 (0.83, 8.02) |
|  | Mask | 1.66 (0.53, 5.22) |

(\*) Obtained from multilevel negative binomial regression analyses

### Relative Risk of Singing with a Face Mask compared to Speaking or Exhaling

In Table 3 each task is compared to a baseline task, along with a corresponding confidence interval. The note task was chosen as the reference. There was a statistically significant difference between droplet numbers for all tasks. The number of droplets was lowest for the note task and highest for exhaling and saying ‘hello’. Most importantly, neither singing a note or ‘Happy Birthday’ transmitted a higher mean droplet number than exhaling or speaking. The decreased likelihood of transmitting droplets when speaking or exhaling when wearing masks is also demonstrated in Figure 2B.

**Table 3:**
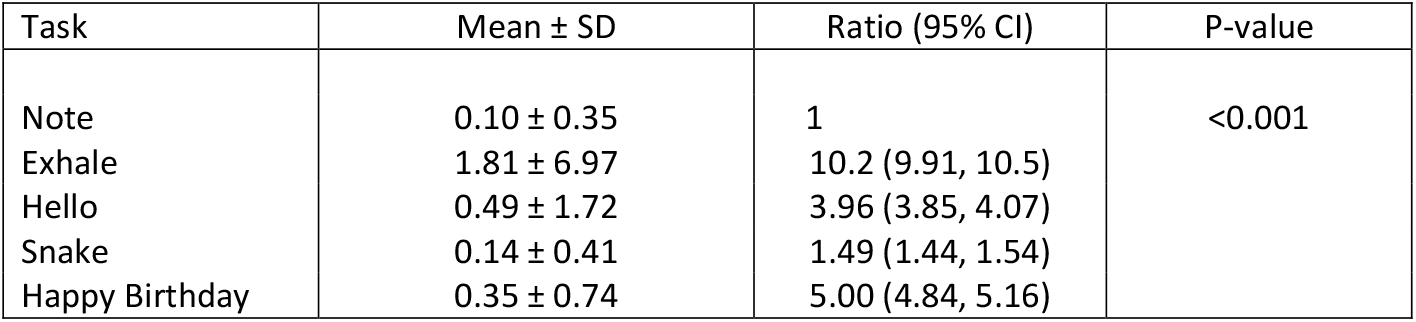
Comparison of mean droplet density across all tasks when wearing a face mask

| Task | Mean ± SD | Ratio (95% CI) | P-value |
| --- | --- | --- | --- |
| Note | 0.10 ± 0.35 | 1 | <0.001 |
| Exhale | 1.81 ± 6.97 | 10.2 (9.91, 10.5) | |
| Hello | 0.49 ± 1.72 | 3.96 (3.85, 4.07) | |
| Snake | 0.14 ± 0.41 | 1.49 (1.44, 1.54) | |
| Happy Birthday | 0.35 ± 0.74 | 5.00 (4.84, 5.16) | |

### Are there ‘Super Emitters?’

This objective was examined graphically in Figure 5. The mean number of droplets per patient for each task was calculated. The objective here was to assess whether some participants always transmitted more droplets than others. Data from the ‘Happy Birthday’ task was not considered as the 7 participants who did this task did not complete the other tasks. Droplets were transmitted relatively consistently between tasks for all participants but three generated dramatically more droplets for singing a note than the others (see also figure 2B). These large variations were abolished when participants wore face masks. There was no consistency in which tasks generated the highest numbers of droplets for individual participants.

**Figure 5:**
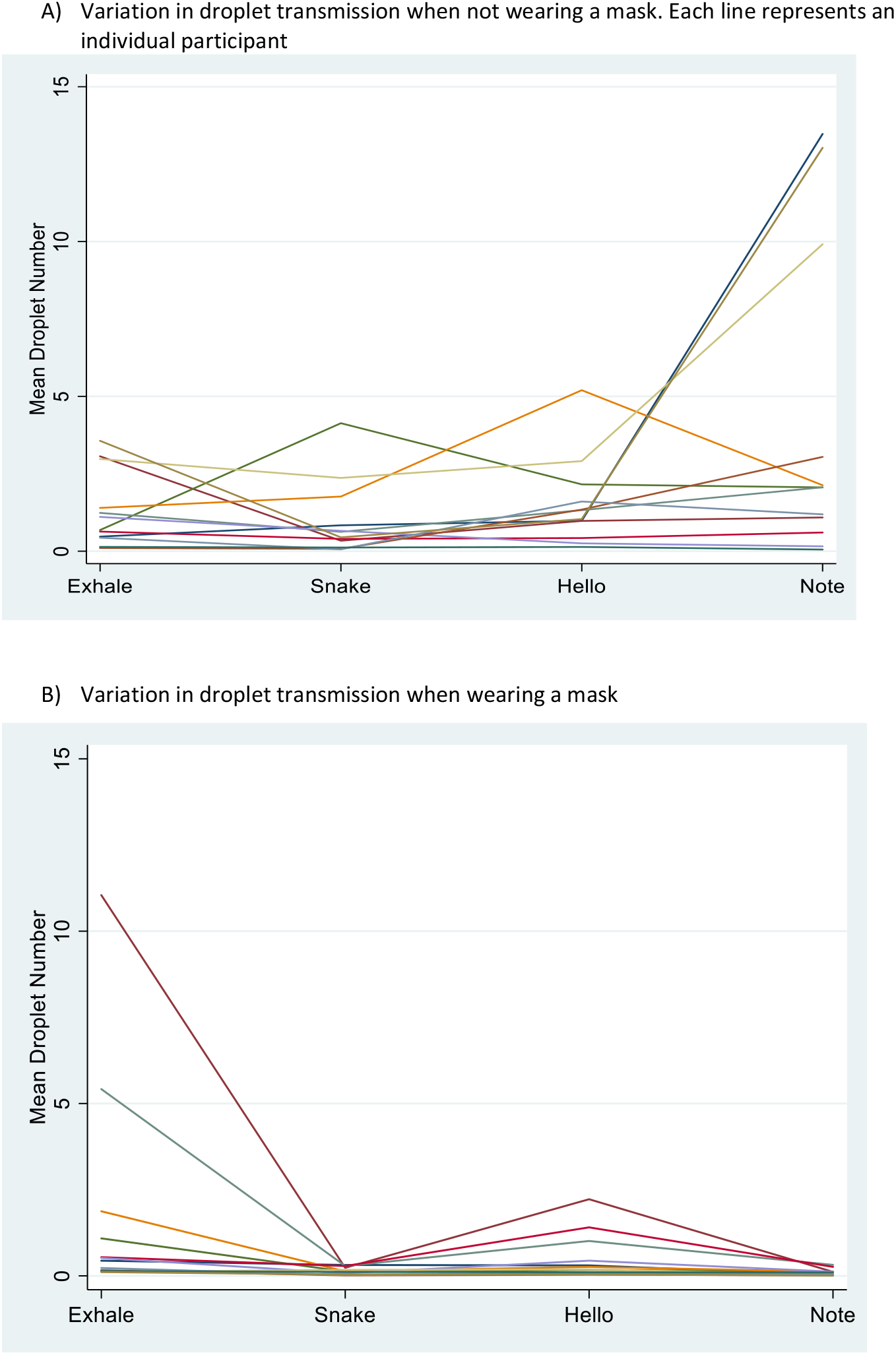
The mean droplet density for each task is plotted showing the differences between individuals

### Correlation with Participant Physical Characteristics

We also assessed whether height, weight, body mass index (BMI) or ethnic background of subjects affected droplet parameters. No consistent findings were observed.

## Discussion

Our data offer novel insights into the spatio-temporal evolution of droplets with and without surgical face masks for 20 subjects who completed an array of verbal tasks including singing. Exhaling produced less droplets than saying ‘hello’ but more than saying ‘snake’, both at 70 dB. Singing a note produced the most droplets whereas singing ‘Happy Birthday’ produced a similar number to speaking despite singing tasks being undertaken at 85 dB. It has been noted that people speak louder when wearing masks.^28^ We did not observe this, but we did show a very striking variation between individuals, with some producing almost no droplets, and others producing large numbers, particularly when singing. There was also no consistency between participants as to which task generated the most droplets. This confirms previous studies which have shown that different people emit more droplets for different tasks and suggests that emission may be highly person and task specific.^11,12^ Crucially, we demonstrated a dramatic reduction in droplet transmission when face masks were worn and inter-individual variations were abolished. Significant transmission of droplets through the face masks occurred only with exhaling and saying ‘hello’. Finally, when wearing a face mask, less droplets were transmitted when singing compared to exhaling or speaking. We did not investigate the size of droplets, nor the impact of evaporation, which have been researched extensively by others.^12,13,15,17^

Our results are consistent with other recent work. Alsved et al demonstrated in a cohort of 12 volunteers that normal singing produced more droplets compared to speech using a combination of a sampling funnel fitted around the participant’s face and light scattering spectroscopy in a subset of 5 participants.^21^ In a UK cohort of 25 professional singers, wide variation was found in droplet transmission across multiple vocal tasks.^12^ Although singing produced more droplets, differences seen were more likely to be due to volume rather than task. The same phenomenon was reported by others.^11,21^ We did not control for this so the higher droplet numbers we showed with singing, which was also louder, may have been due to either of these phenomena. We found no sex-related differences, in line with Gregson et al.^12^ Our results clearly show that inter-individual variation in droplet transmission is very wide. There are at least fifty-fold differences in the maximum and time averaged droplet numbers imaged. Duration of droplet production can range from 10-2000 milliseconds. We were unable to identify any specific characteristics that predicted these variations. Nonetheless our findings may significantly influence future modelling of aerosol transmission and infectivity of SARS-CoV-2.^29^

Our findings also concur with observational real-world data regarding the value of face masks. A recent meta-analysis demonstrated that face masks could lead to a large reduction of risk of SARS-CoV-2 disease transmission (adjusted odds ratio 0.15, 95%CI: 0.07-0.34).^30^ It makes sense,. therefore, that surgical face masks reduce the transmission of coronaviruses.^20^ Laboratory based experiments have similarly demonstrated efficacy, often with at least 80% reduction.^28,31,32^ Our data additionally demonstrate that wearing a face mask dramatically reduces the inter-individual variability of droplet transmission while speaking and singing. Asadi et al. demonstrated that wearing a face mask would reduce droplet load from so-called ‘super-emitters’ and offer some protection to disease spread.^28^ Our data would strongly support this supposition.

We did not examine the amount of leakage around the sides of the masks. Viola et al. did this and demonstrated significant leakage jets that may present major hazards.^31^ But their study examined coughing where droplets were 50 μm in diameter and peak velocity was 8 m/s despite wearing masks. Verma et al. also explored this for coughs and showed only minor escape around the top of masks and at low velocities.^32^ For speaking or singing, droplets are smaller^11^ and the velocity without masks in our study was less than 1 m/s. When face masks were worn, transmission velocity was less than 10% of this. Even if there was leakage, it would be at a similar slow velocity which would lead to very slow diffusion of the aerosols away from the speaker or singer.^29,32^ SARS-CoV-2 becomes less infectious with time, so even if there is significant mask leakage, the slow diffusion rate will decrease the risk of seeding infection, particularly in a place of worship that stringently enforces social distancing with a short service.^10^ But it strengthens the case for using masks which can actively kill virus such as those infused with copper or zinc.^33,34^

### Wider Implications

Consensus opinion has previously been that it would not be safe for singers to rehearse together unless there was a COVID-19 vaccine available and a 95% effective treatment in place.^35^ In addition, singing can be an emotive topic so good quality data is needed. Based on the results of this study, we conclude that by wearing face masks, the risks of disease transmission when singing indoors can be reduced to those of sitting quietly or speaking normally. This makes singing no more likely to transmit virus in a communal worship setting. There are benefits of singing, most notably with mental health. An example is the sound of singing in Italian cities, which was used to boost national morale during the first national lockdown.^36^ Combined with other mitigation strategies currently recommended such as social distancing, increasing ventilation and reducing volume, we believe it is reasonable for congregational singing to return to places of worship which have remained open during lockdowns in the UK.^7^ Indeed, our questionnaire of 1000 worshippers also demonstrates a longing to return to singing, even if it means wearing a face mask and worshippers all report that their places of worship strongly enforce all the government guidelines including social distancing.^10^ Allowing congregational singing indoors could vastly improve congregants’ worshipping experiences, and restore ‘a sense of celebration.’^8^

### Limitations

Our study has several limitations. Firstly, it was not possible to determine the exact lower limit of particle diameter that can be detected by our apparatus. However, our validation test using a medical nebuliser demonstrated that our method could detect droplets which are on average 3 µm in size. The range from 3-5 µm is the key droplet size for disease transmission and we detect this. Our study utilised only surgical type IIR face masks, and hence would not be applicable to all face coverings. In particular, some homemade masks are less effective in reducing droplet transmission compared to surgical face masks.^32,37,38^ Congregants in places of worship may therefore also need to be provided with additional guidance regarding the type of face coverings to wear. Finally, we only assessed individuals singing in a controlled laboratory. There are additional considerations in a real world setting such as in a place of worship which need to be considered, including ventilation and spacing of congregants.

## Conclusions

Our work explored how the aerosol plume evolves as function of space and time and looked at the efficiency of masks with vocal activities. Using high-resolution imaging, we demonstrated the wide variation in droplet transmission with different vocal tasks. Face masks eliminate this variation and are efficacious in reducing droplet spread when singing by reducing transmission as well as velocity of droplets when egress occurs. Face masks could potentially be used alongside other COVID-19 mitigation measures to allow for singing indoors. Our results add to the evidence that supports relaxation of guidance regarding indoor singing.

## Data Availability

Data from the study is available on request from the corresponding author. This may be subject to a data access request agreement.

## Acknowledgments

We would like to thank all the volunteers who participated in these experiments. We acknowledge partial support from Wellcome/EPSRC Centre for Interventional and Surgical Sciences (WEISS) (203145Z/16/Z). LBL is supported by the National Institute for Health Research University College London (UCL) Hospitals Biomedical Research Centre. MKT acknowledges the Royal Society Wolfson Fellowship and the NICEDROPS project supported by the European Research Council (ERC) under the European Union’s Horizon 2020 research and innovation programme under grant agreement no. 714712.

## Role of funding source

None

## Competing interests

The authors declare no competing interests

## Supplementary Materials

Supplementary Video 1: Video of time vector plots of droplets being produced, after a participant sang the note ‘la’.

**Supplementary Table 1:** Demographic data of participants

|  |  |
| --- | --- |
|  | N=20 |
| Age median (IQR) | 42.0 (27.0) |
| Female n( %) | 14 (70) |
| Ethnicity n (%) |  |
| White British | 11 (55) |
| Any Other White Background | 8 (40) |
| Asian/Asian British | 1 (5) |
| Body Mass Index n (%) |  |
| 18.5 to <25 | 11 (55) |
| 25 to <30 | 6 (30) |
| ≥30 | 1 (5) |
| Missing | 2 (10) |
| Smoking Status |  |
| Non-smoker | 18 (90) |
| Ex-smoker | 2 (10) |
| Alcohol Intake |  |
| None | 8 (40) |
| Small (1 small drink per day/a couple of pints of beer a week) | 4 (20) |
| Average (1 medium drink per day/up to 7 pints of beer a week) | 5 (25) |
| Large: (1 large drink per day/more than 7 pints of beer a week) | 3 (15) |

**Supplementary Figure 1.**
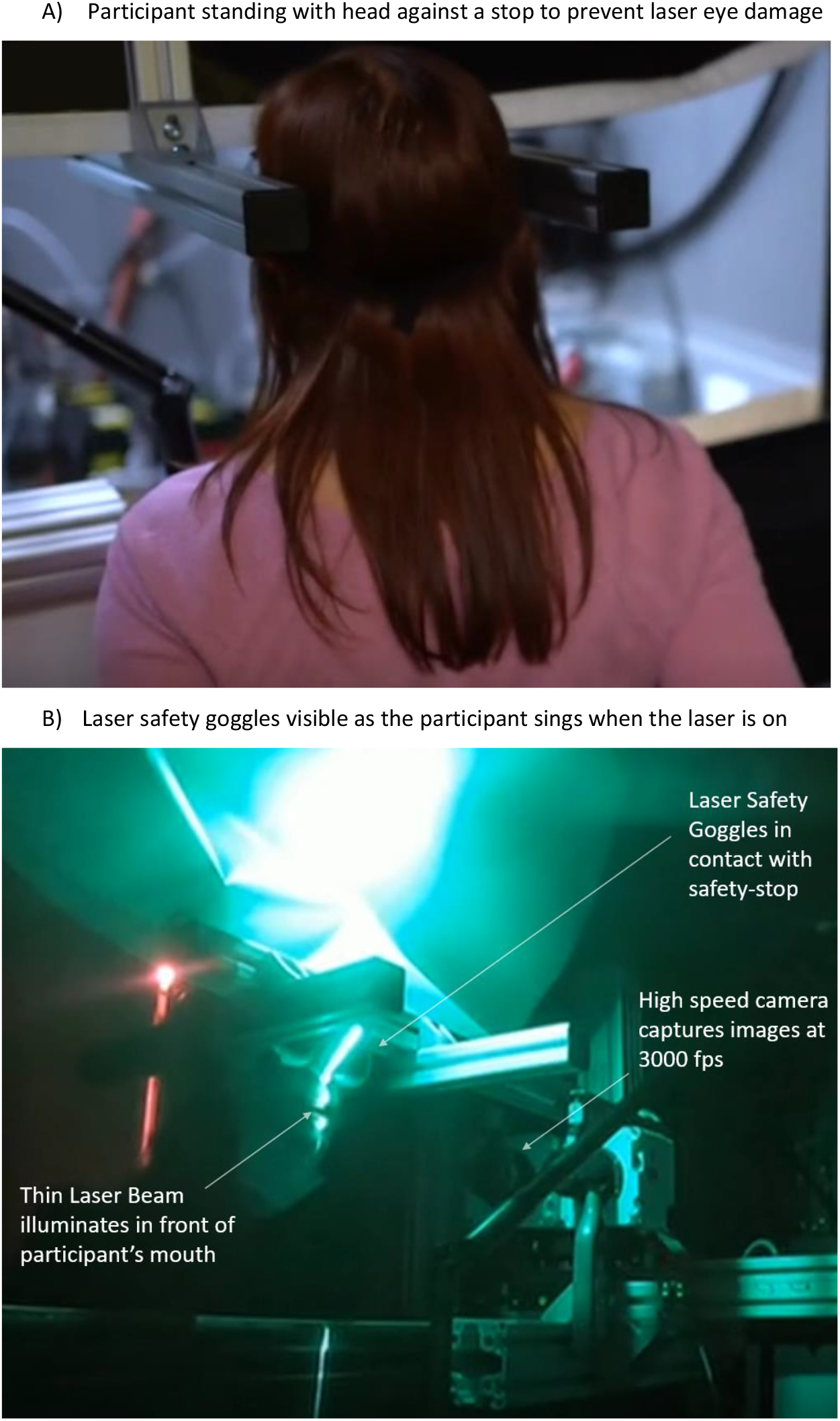

**Supplementary Figure 2:**
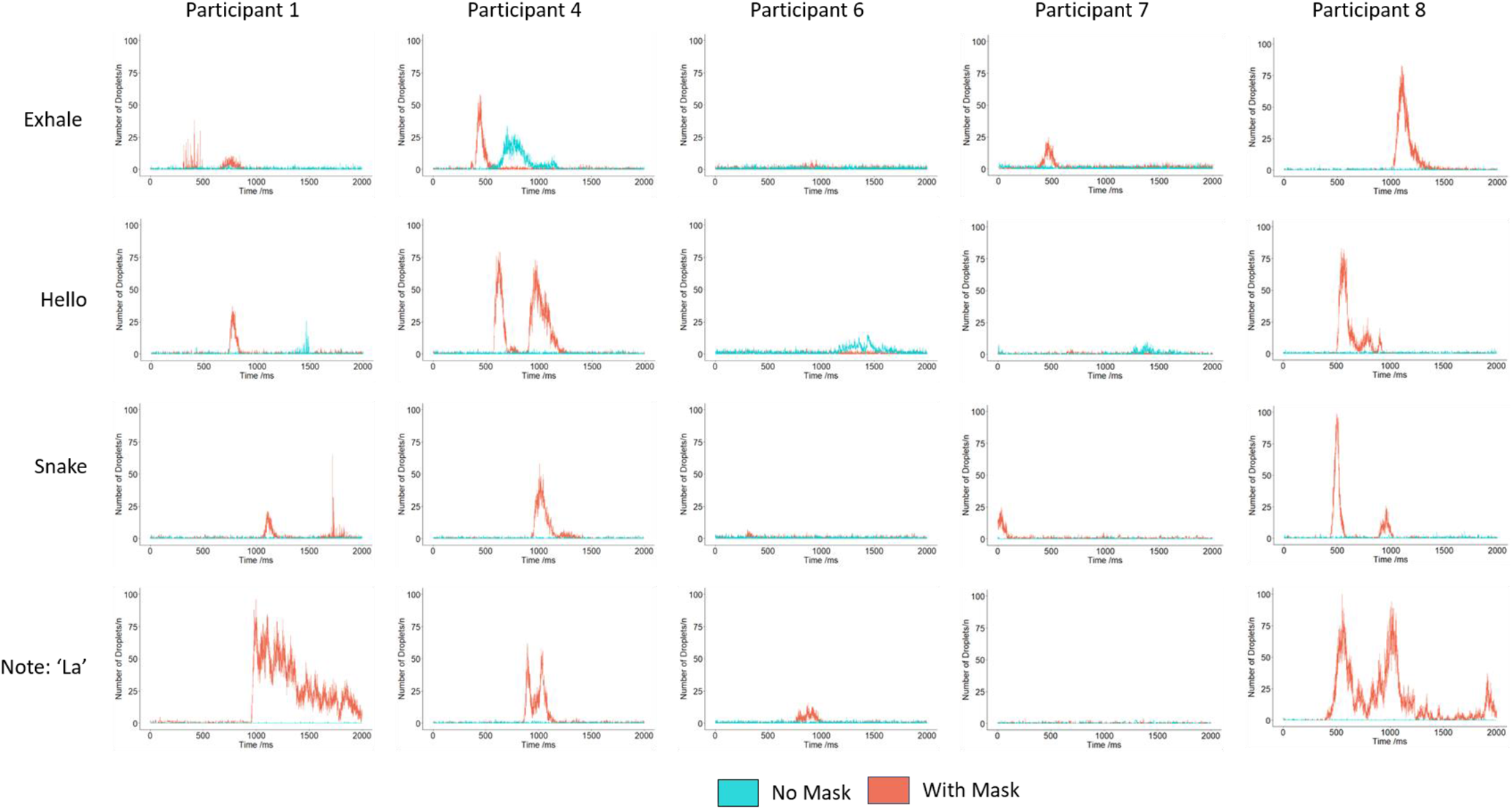
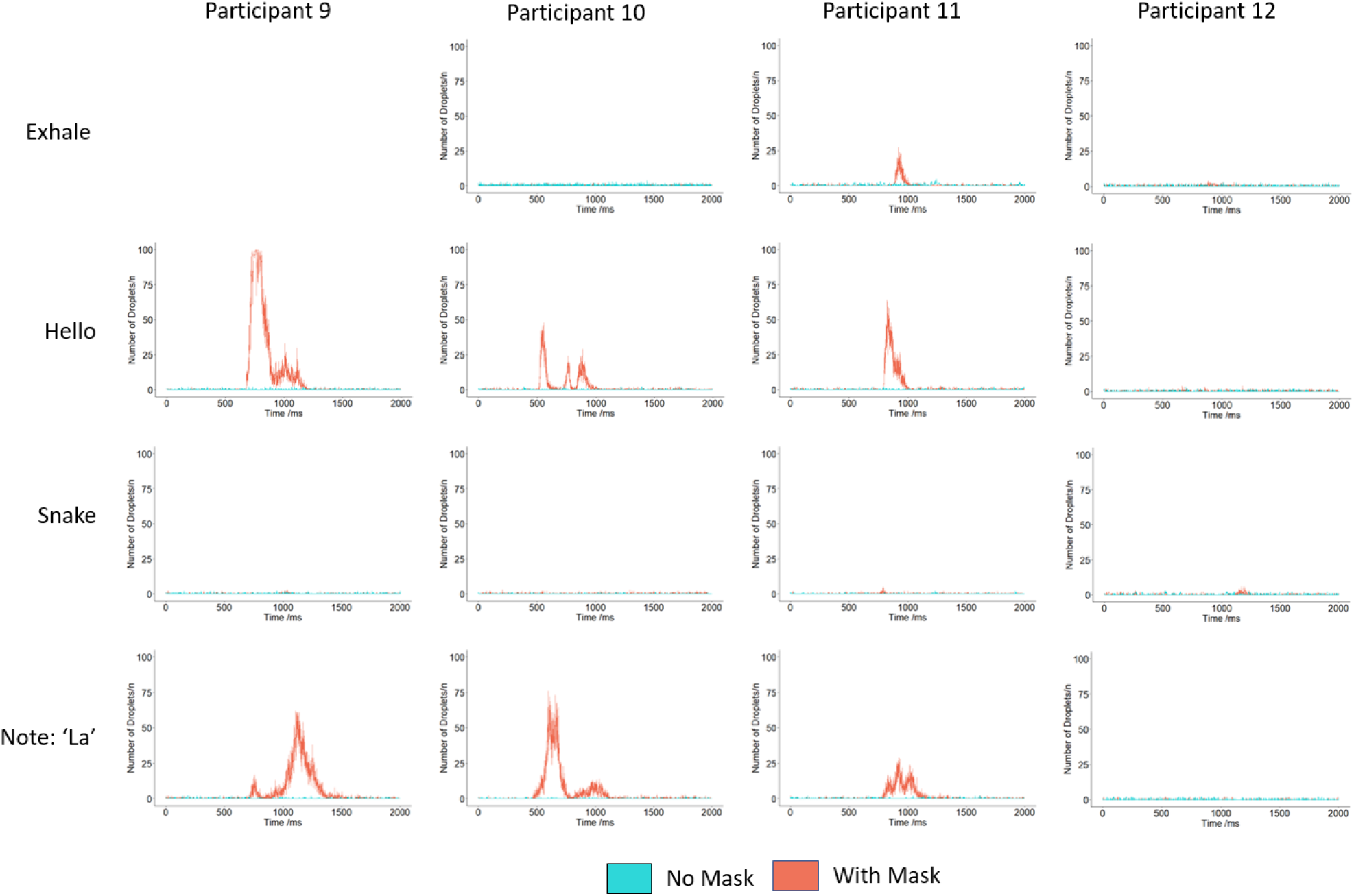
Time resolved transmission of droplets for all other participants not represented in Figure 2B, when exhaling; saying ‘hello’; saying ‘snake’ and singing the note ‘la’. Again, there was no consistent pattern for number of droplets produced across the various tasks. No result is available for participant 9 for exhaling as the laser was accidentally knocked creating erroneous results and is hence omitted.

**Supplementary Figure 3:**
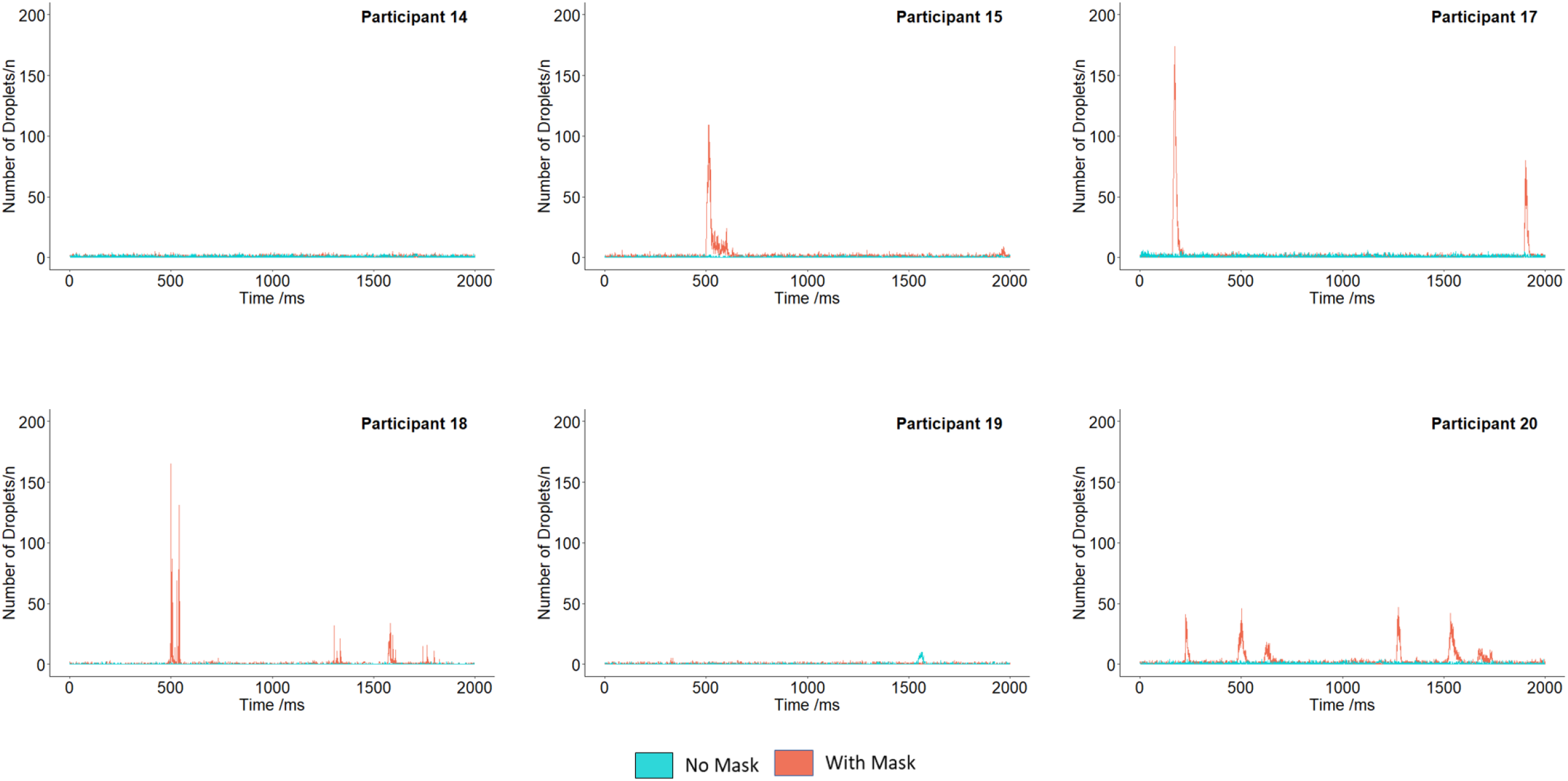
Time resolved transmission of droplets for 6 participants who sang the first two lines of ‘Happy Birthday’. There is no consistent pattern between individuals as noted with the other tasks. No result is available for participant 16 as the laser was accidentally knocked creating erroneous results and is hence omitted.

## Notes

### Competing Interest Statement

The authors have declared no competing interest.

### Funding Statement

Grant funding from Wellcome Trust and EPSRC through the Wellcome / EPSRC Centre for Interventional and Surgical Sciences (WEISS) (203145Z/16/Z). LBL is supported by the National Institute for Health Research University College London (UCL) Hospitals Biomedical Research Centre. MKT acknowledges the Royal Society Wolfson Fellowship and the NICEDROPS project supported by the European Research Council (ERC) under the European Union's Horizon 2020 research and innovation programme under grant agreement no. 714712.

## References

1. Stadnytskyi, V., Anfinrud, P. & Bax, A. Breathing, speaking, coughing or sneezing: What drives transmission of SARS-CoV-2? J. Intern. Med. (2021). doi:10.1111/joim.13326

2. Public Health England. How to stop the spread of coronavirus (COVID-19). www.gov.uk (2021). Available at: https://www.gov.uk/government/publications/how-to-stop-the-spread-of-coronavirus-covid-19/how-to-stop-the-spread-of-coronavirus-covid-19. (Accessed: 21st April 2021)

3. Centres for Disease Control and Prevention. Considerations for Events and Gatherings. (2021). Available at: https://www.cdc.gov/coronavirus/2019-ncov/community/large-events/considerations-for-events-gatherings.html. (Accessed: 20th April 2021)

4. Kim, S. et al. Evaluation of COVID-19 epidemic outbreak caused by temporal contact-increase in South Korea. Int. J. Infect. Dis. 96, 454–457 (2020).

5. Mat, N. F. C., Edinur, H. A., Razab, M. K. A. A. & Safuan, S. A single mass gathering resulted in massive transmission of COVID-19 infections in Malaysia with further international spread. J. Travel Med. 27, 1–4 (2020).

6. Hamner, L. et al. High SARS-CoV-2 Attack Rate Following Exposure at a Choir Practice — Skagit County, Washington, March 2020. MMWR. Morb. Mortal. Wkly. Rep. 69, 606–610 (2020).

7. Public Health England. Covid-19: suggested principles of safer singing. (2020). Available at: https://www.gov.uk/government/publications/covid-19-suggested-principles-of-safer-singing/covid-19-suggested-principles-of-safer-singing. (Accessed: 20th April 2021)

8. Burgess, K. Hark! Tomorrow heralds early return of singing. The Times (2021).

9. Ministry of Housing, Communities & Local Government. COVID-19: guidance for the safe use of places of worship during the pandemic from 4 July. (2021). Available at: https://www.gov.uk/government/publications/covid-19-guidance-for-the-safe-use-of-places-of-worship-during-the-pandemic-from-4-july. (Accessed: 20th April 2021)

10. Ho, K. M. A. et al. Face mask acceptability for communal religious worship during the COVID-19 pandemic: results from the CONFESS study. J. Relig. Health 60, in press (2021).

11. Asadi, S. et al. Aerosol emission and superemission during human speech increase with voice loudness. Sci. Rep. 9, (2019).

12. Gregson, F. K. A. et al. Comparing aerosol concentrations and particle size distributions generated by singing, speaking and breathing. Aerosol Sci. Technol. 55, 681–691 (2021).

13. Johnson, G. R. et al. Modality of human expired aerosol size distributions. J. Aerosol Sci. 42, 839–851 (2011).

14. Anderson, E. L., Turnham, P., Griffin, J. R. & Clarke, C. C. Consideration of the Aerosol Transmission for COVID-19 and Public Health. Risk Anal. 40, (2020).

15. Walker, J. S., Gregson, F. K. A., Michel, S. E. S., Bzdek, B. R. & Reid, J. P. Accurate Representations of the Microphysical Processes Occurring during the Transport of Exhaled Aerosols and Droplets. Cite This ACS Cent. Sci (2021). doi:10.1021/acscentsci.0c01522

16. Prather, K. A. et al. Airborne transmission of SARS-CoV-2. Science 370, 303–304 (2020).

17. Morawska, L. et al. Size distribution and sites of origin of droplets expelled from the human respiratory tract during expiratory activities. J. Aerosol Sci. 40, 256–269 (2009).

18. Robinson, J. F. et al. Efficacy of face coverings in reducing transmission of COVID-19: calculations based on models of droplet capture. 1–7 (2020).

19. Anand, S. & Mayya, Y. S. Size distribution of virus laden droplets from expiratory ejecta of infected subjects. Sci. Rep. 10, 1–9 (2020).

20. Leung, N. H. L. et al. Respiratory virus shedding in exhaled breath and efficacy of face masks. Nat. Med. 26, 676–680 (2020).

21. Alsved, M. et al. Exhaled respiratory particles during singing and talking. Aerosol Science and Technology 54, 1245–1248 (2020).

22. Echternach, M. et al. Impulse dispersion of aerosols during singing and speaking: A potential COVID-19 transmission pathway. Am. J. Respir. Crit. Care Med. 202, 1584–1587 (2020).

23. Asadi, S. et al. Effect of voicing and articulation manner on aerosol particle emission during human speech. PLoS One 15, 1–15 (2020).

24. Bahl, P., De Silva, C., Chughtai, A. A., MacIntyre, C. R. & Doolan, C. An experimental framework to capture the flow dynamics of droplets expelled by a sneeze (Manuscript under review). Exp. Fluids Manuscript Under Review (2020). doi:10.1007/s00348-020-03008-3

25. Bahl, P. et al. Face coverings and mask to minimise droplet dispersion and aerosolisation : a video case study. 1–2 (2020). doi:10.1136/thoraxjnl-2020-215748

26. Schraer, R. Covid: Faith groups’ singing studied for coronavirus risk. BBC News (2020). Available at: https://www.bbc.co.uk/news/health-54390549. (Accessed: 20th April 2021)

27. Covid-19: PM announces four-week England lockdown - BBC News.

28. Asadi, S. et al. Efficacy of masks and face coverings in controlling outward aerosol particle emission from expiratory activities. Sci. Rep. 10, 1–13 (2020).

29. Vuorinen, V. et al. Modelling aerosol transport and virus exposure with numerical simulations in relation to SARS-CoV-2 transmission by inhalation indoors. Saf. Sci. 130, 1–23 (2020).

30. Chu, D. K. et al. Physical distancing, face masks, and eye protection to prevent person-to-person transmission of SARS-CoV-2 and COVID-19: a systematic review and meta-analysis. Lancet 395, 1973–1987 (2020).

31. Viola, I. M. et al. Face Coverings, Aerosol Dispersion and Mitigation of Virus Transmission Risk. IEEE Open J. Eng. Med. Biol. 2, 26–35 (2021).

32. Verma, S., Dhanak, M. & Frankenfield, J. Visualizing the effectiveness of face masks in obstructing respiratory jets. Phys. Fluids 32, 61708 (2020).

33. Jung, S. et al. Copper-coated polypropylene filter face mask with SARS-COV-2 antiviral ability. Polymers (Basel). 13, 1–10 (2021).

34. Gopal, V. et al. Zinc-embedded fabrics inactivate SARS-CoV-2 and influenza A virus. bioRxiv Prepr. Serv. Biol. 1–27 (2020). doi:10.1101/2020.11.02.365833

35. Naunheim, M. R. et al. Safer Singing During the SARS-CoV-2 Pandemic: What We Know and What We Don’t. J. Voice (2020). doi:10.1016/j.jvoice.2020.06.028

36. Corvo, E. & De Caro, W. COVID-19 and Spontaneous Singing to Decrease Loneliness, Improve Cohesion, and Mental Well-Being: An Italian Experience. Psychol. Trauma Theory, Res. Pract. Policy 12, 247–248 (2020).

37. Fischer, E. P. et al. Low-cost measurement of face mask efficacy for filtering expelled droplets during speech. Sci. Adv 6, (2020).

38. Kähler, C. J. & Hain, R. Fundamental protective mechanisms of face masks against droplet infections. J. Aerosol Sci. 148, 105617 (2020).

